# Predictors for participation in DNA self-sampling of childhood cancer survivors in Switzerland

**DOI:** 10.1101/2021.05.25.21257796

**Authors:** Nicolas Waespe, Sven Strebel, Denis Marino, Veneranda Mattiello, Fanny Muet, Tiago Nava, Christina Schindera, Fabien N. Belle, Luzius Mader, Adrian Spoerri, Claudia E. Kuehni, Marc Ansari

**Affiliations:** Childhood Cancer Research Group, Institute of Social and Preventive Medicine, University of Bern, Bern, Switzerland; CANSEARCH research platform for paediatric oncology and haematology, Faculty of Medicine, Department of Pediatrics, Gynecology and Obstetrics, of the University of Geneva, Geneva, Switzerland; Graduate School for Cellular and Biomedical Sciences (GCB), University of Bern, Bern, Switzerland; Graduate School for Health Sciences (GHS), University of Bern, Bern, Switzerland; Division of Pediatric Oncology and Hematology, Department of Women, Children, and Adolescents, University Hospital of Geneva, Geneva, Switzerland; Pediatric Oncology/Hematology, University Children’s Hospital Basel, University of Basel, Basel, Switzerland; Center for Primary Care and Public Health (Unisanté), Institute of Social and Preventive Medicine (IUMSP), University of Lausanne, Lausanne, Switzerland; SwissRDL - Medical Registries and Data Linkage, Institute of Social and Preventive Medicine, University of Bern, Bern, Switzerland; Division of Pediatric Oncology and Hematology, Department of Pediatrics, University Hospital of Bern, Bern, Switzerland

**Author notes:** **Corresponding author:** Prof. Dr. med. Marc Ansari, MD; Division of Pediatric Oncology and Hematology, Department of Women, Children, and Adolescents, University Hospital of Geneva, Rue Willy-Donzé 6; Bureau 5-507; 1211 Genève, Switzerland; F. Shared equally contributing senior authors (Claudia E. Kuehni, Marc Ansari).

**Keywords:** childhood cancer, cancer survivors, DNA, cohort study, drug side effects, second primary neoplasm, genetic predisposition, registry, genetic testing, genetic polymorphism

## Abstract

Research on germline genetic variants relies on a sufficient number of eligible participants which is difficult to achieve for rare diseases such as childhood cancer. With self-collection kits using saliva or buccal swabs, participants can contribute genetic samples conveniently from their home. We identified determinants of participation in DNA self-collection in this cross-sectional study.

We invited 928 childhood cancer survivors in Switzerland with a median age of 26.5 years (interquartile range 18.8-36.5), of which 463 (50%) participated. Foreign nationality (odds ratio [OR] 0.5, 95%-confidence interval [CI] 0.4-0.7), survivors aged 30-39 years at study versus other age groups (OR 0.5, CI 0.4-0.8), and those with a known cancer predisposition syndrome (OR 0.5, CI 0.3-1.0) participated less. Survivors with a second primary neoplasm (OR 1.9, CI 1.0-3.8) or those living in a French or Italian speaking region (OR 1.3, 1.0-1.8) tended to participate more.

We showed that half of survivors participate in germline DNA self-sampling relying completely on mailing of sample kits. Foreign nationality, age 30-39 years, and cancer predisposition syndromes were associated with less participation. More targeted recruitment strategies may be advocated for these subgroups. To increase participation in DNA self-sampling, understanding and perceptions of survivors need to be better understood.

## 1. INTRODUCTION

Cancer survivorship and associated health complications have become increasingly important with improved childhood cancer survival.[1] Chronic conditions like cardiac, pulmonary, auditory, endocrine, reproductive, and neurocognitive complications, and second primary neoplasms (SPNs) are gaining importance in research and clinical care.[2] By age 50 years, childhood cancer survivors have twice as many severe chronic health conditions compared to controls.[3] Mortality in survivors is more than 10-times higher than in the general population.[4] While many demographic, clinical, and treatment-related risk factors are known, the contribution of genetic variation in the development of health complications is still poorly understood. Therefore, research group collect germline DNA samples systematically to enable research on genetic risk factors.[5–7] Self-sampling of germline DNA by participants has increased the reach of sample collection because many do not attend regular medical care. Since the 1990’s buccal swabs are used. In the 2010’s saliva collection kits became widely available which are easy to use and yield DNA of good quality and sufficient quantity.[8] Saliva collection kits do not need time-sensitive processing or cooling like blood, but can be collected, transported, and stored in ambient conditions for years. Collection of saliva samples by participants themselves is feasible and effective.[5] Self-collection may, however, be affected by participation bias. Ness et al. identified female sex, white race/ethnicity, college graduation, never smoking, accessing the healthcare system in the past 2 years, and having a second malignant neoplasm as predictors for participating in saliva sample self-collection.[5] Outside of the US, predictors for participation in childhood cancer survivors have not been studied. They might differ based on the perception of the healthcare system and acceptance of genetic research. We assessed the response rate and investigated predictors for participation in saliva self-sampling in Switzerland as part of the national Germline DNA Biobanking for Childhood Cancer and Blood Disorders (BISKIDS) project.

## 2. METHODS

### 2.1. Study design

We used contact information from the Swiss Childhood Cancer Registry (SCCR) at the Institute of Social and Preventive Medicine, University of Bern, Switzerland to invite childhood cancer survivors for germline DNA self-collection in this cross-sectional study design. Addresses were collected from hospitals involved in primary and follow-up care and updated through the Swiss national postal service.

We obtained data on patient characteristics from the SCCR. In September 2019, participants received self-sampling kits by postal mail at their home and were asked to send them to the germline DNA Biobank Switzerland for Childhood Cancer and Blood Disorders (BISKIDS), which is part of the Paediatric Biobank for Research in Haematology and Oncology (BaHOP) Geneva, Switzerland. BaHOP was awarded a VITA label from the Swiss Biobanking Platform (SBP; www.biobanksqan.ch/#/biobanks/3919; bbmrieric:ID:CH_HopitauxUniversitairesGeneve:collection:CH_BaHOP). All participants received an informed consent form together with the saliva kit. The Geneva Cantonal Commission for Research Ethics has approved the BaHOP biobank (approval PB_2017-00533) and the associated “Genetic Risks for Childhood Cancer Complications Switzerland (GECCOS)” study, which will utilize the samples (approval 2020-01723).

### 2.2. Study population

Eligible for invitation to our study were participants who were: (i) registered in SCCR; (ii) Swiss residents; (iii) treated in one of nine paediatric hospitals caring for children with cancers; (iv) diagnosed with a neoplasm according to the International Classification of Childhood Cancers, 3^rd^ edition (ICCC-3), or Langerhans cell histiocytosis before age 21 years from 1976 to 2017; (v) exposed to lung toxic (chest radiotherapy) or ototoxic treatment (brain radiotherapy with ≥30 Gray or platinum chemotherapy); and (vi) survivors of 2 years or more since childhood cancer diagnosis as of July 2019. We excluded participants who: (I) had declined participation in research projects; (II) had died; or (III) did not have a valid address in Switzerland.

### 2.3. Outcome definition and clinical characteristics

Our main outcome was participation in the germline DNA sample collection, defined as returning the DNA sample and the signed consent form. Non-participation was defined as active decline or non-response until December 2020 (end of follow-up). Clinical information was extracted from the SCCR. We recorded the age at first neoplasm in 5-year groups, calendar year at first neoplasm diagnosis and age at survey in 10-year groups, and the ICCC-3 main category [9] of the first neoplasm. Chemotherapy, radiotherapy, and haematopoietic stem cell transplantation were classified as “yes” if given during the first neoplasm treatment. We classified cancer predisposition syndrome and second primary neoplasms as previously described.[10]

### 2.4. Sample collection

Survivors received by postal mail a letter with information on the planned DNA sample collection, and a form with which they could opt and a prepaid return envelope. Those who did not opt out received 3 weeks later a parcel including (i) detailed information on the biobanking project and an informed consent form to sign, (ii) one Oragene DNA OG-500 saliva sampling kit (manufactured by DNA Genotek, Ottawa, Ontario, Canada) with material for return by regular mail (liquid-tight bio-specimen bag, bubble wrap), (iii) information on saliva sample collection and shipment provided by the manufacturer (www.dnagenotek.com/row/products/collection-human/oragene-dna/500-series/OG-500.html), (iv) a graphical abstract of the workflow of saliva collection and return of samples and consent forms, and (v) prepaid return envelopes for return of the consent form and the saliva sample. We sent two letters as reminders to those who did not return the sample, one after 6 weeks and another one after 8 weeks.

We offered participants the opportunity to receive additional information through (i) a project specific e-mail address, and (ii) a dedicated telephone hotline. Both were operated by the study coordinator, a trained specialist in paediatric haematology and oncology with experience in cancer genetics, or biobank staff in case of absence.

### 2.5. Statistical analysis

We compared demographic, neoplasm, treatment, and follow-up information of participants with non-participants. Univariable logistic regression and multivariable logistic regression models were fitted to identify determinants of participation. Covariates were kept in the multivariable logistic regression model using backward selection. We removed covariates with p ≥ 0.2. We additionally adjusted for sex and age at first primary neoplasm diagnosis in the model. We used the software Stata version 15 (Stata Corporation, Austin, Texas) for analyses. Statistical uncertainty of estimates was expressed as 95%-confidence intervals.

## 3. RESULTS

### 3.1. Characteristics of cohort

We traced and contacted 928 of 1,215 eligible childhood cancer survivors (**Figure 1**). Of those we contacted, 463 (50%) returned a germline DNA sample. Median age at diagnosis was 8.7 years (interquartile range [IQR] 3-13) and at invitation 26.5 years (IQR 19-37; **Table 1**). The most common diagnoses were central nervous system tumours (28%) and lymphomas (21%) which reflected our selection process of inviting former childhood cancer patients with pulmotoxic and ototoxic treatments. Most patients had been treated with chemotherapy, about two thirds with radiotherapy, and a minority with haematopoietic stem cell transplantation. Relapse had been confirmed in 21% of survivors, a second primary neoplasm in 4% and a cancer predisposition syndrome (CPS) in 5%. Survivors were predominantly from the German (n=633, 68%) language region and the remaining from the French (n=262, 28%) and Italian (33, 4%) language region, which roughly reflects the language distribution in Switzerland.

**Table 1:**
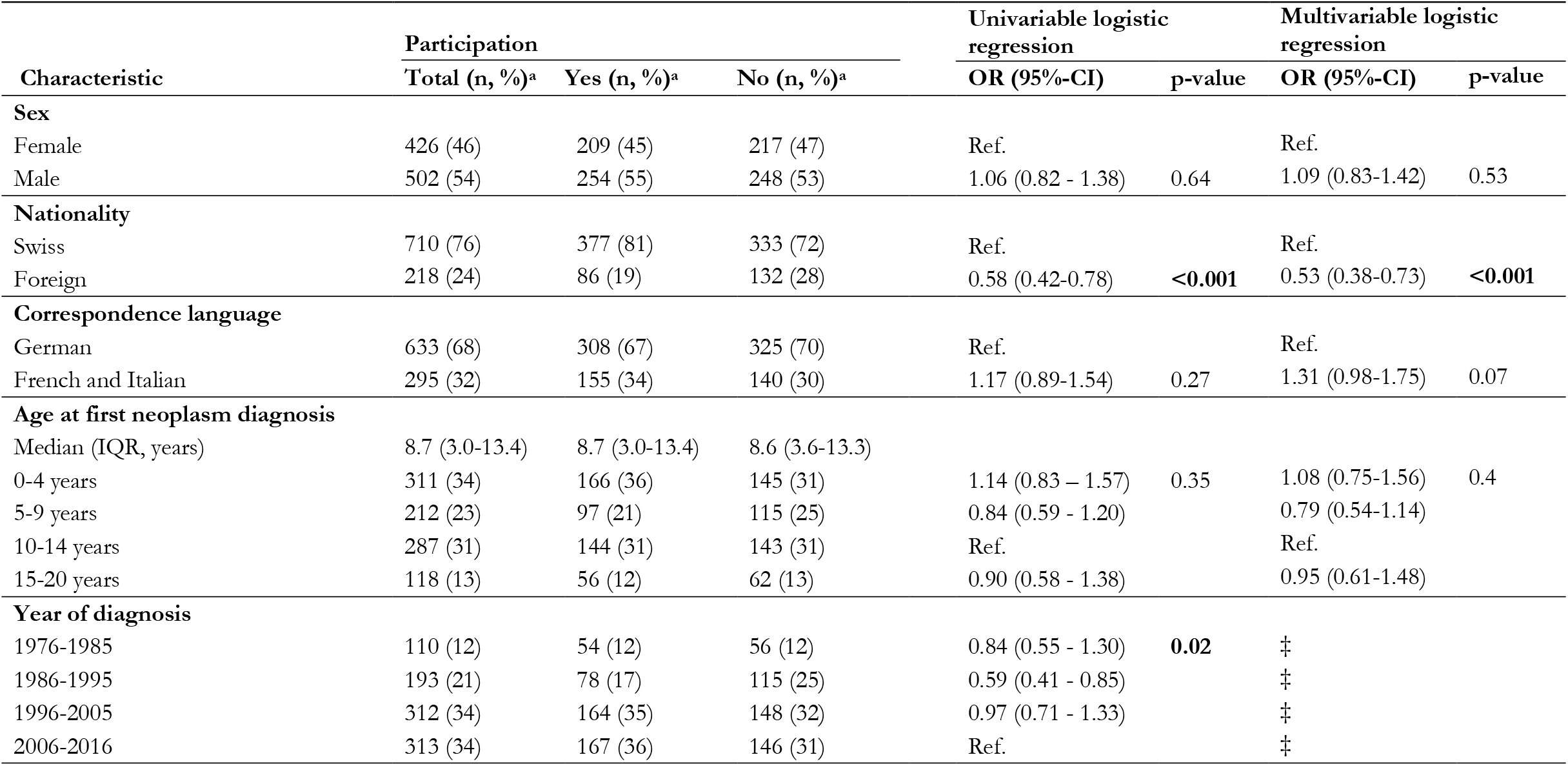
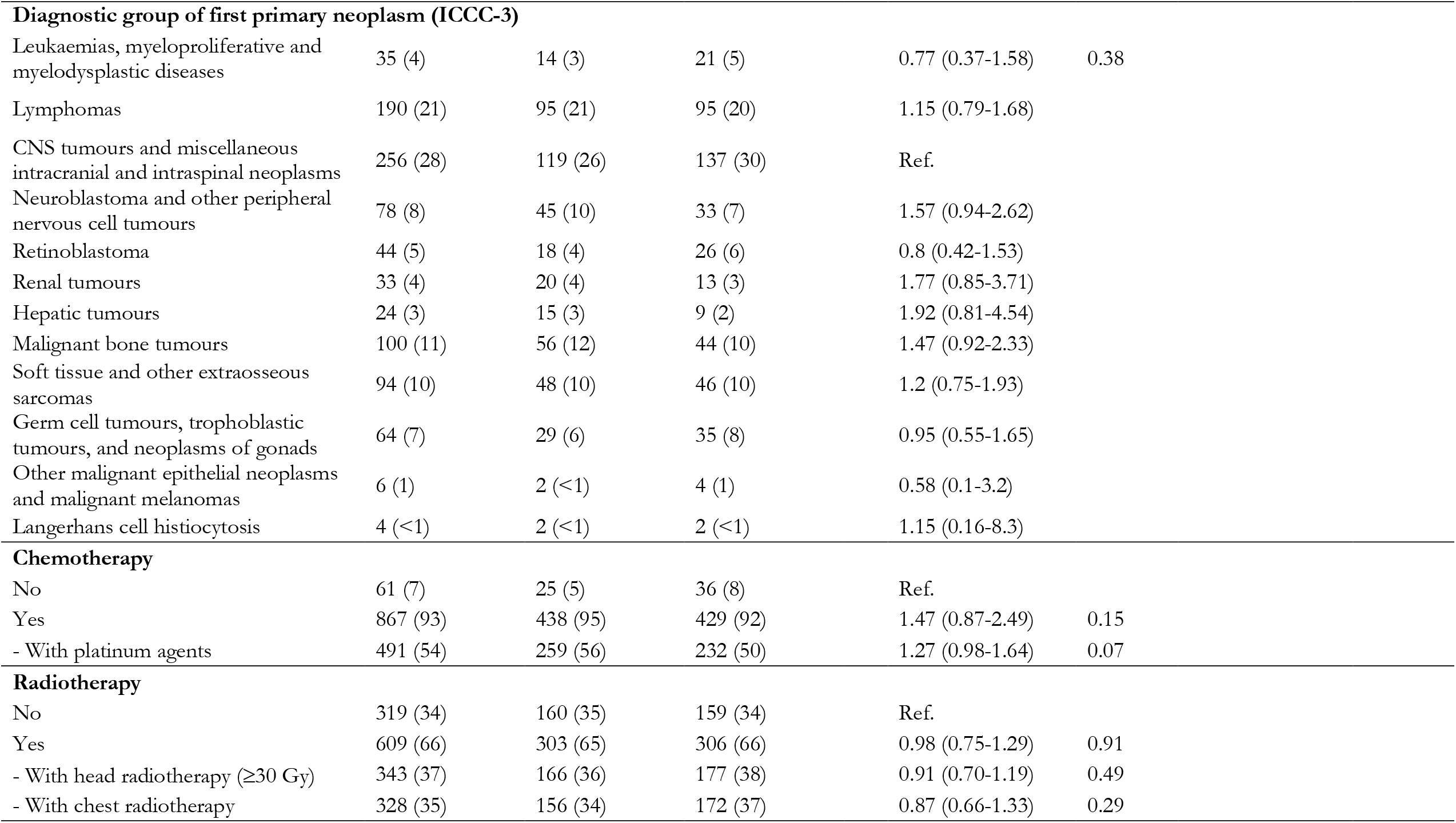
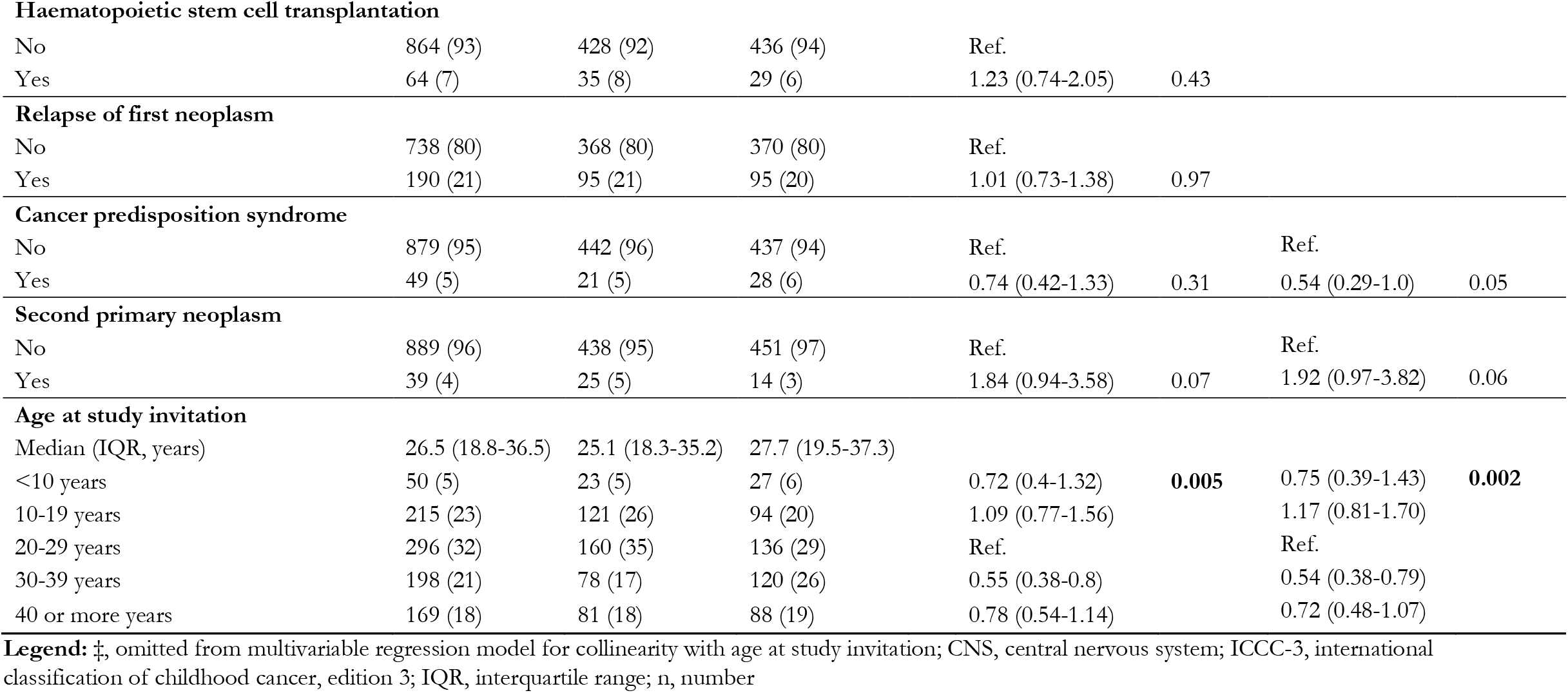
Characteristics of 928 Swiss childhood cancer survivors invited to home collection of germline DNA, stratified by participation status.

| Characteristic | Participation |  |  | Univariable logistic regression |  | Multivariable logistic regression |  |
| --- | --- | --- | --- | --- | --- | --- | --- |
|  | Total (n, %)<sup>a</sup> | Yes (n, %)<sup>a</sup> | No (n, %)<sup>a</sup> | OR (95%-CI) | p-value | OR (95%-CI) | p-value |
| **Sex** |  |  |  |  |  |  |  |
| Female | 426 (46) | 209 (45) | 217 (47) | Ref. |  | Ref. |  |
| Male | 502 (54) | 254 (55) | 248 (53) | 1.06 (0.82 - 1.38) | 0.64 | 1.09 (0.83-1.42) | 0.53 |
| **Nationality** |  |  |  |  |  |  |  |
| Swiss | 710 (76) | 377 (81) | 333 (72) | Ref. |  | Ref. |  |
| Foreign | 218 (24) | 86 (19) | 132 (28) | 0.58 (0.42-0.78) | **<0.001** | 0.53 (0.38-0.73) | **<0.001** |
| **Correspondence language** |  |  |  |  |  |  |  |
| German | 633 (68) | 308 (67) | 325 (70) | Ref. |  | Ref. |  |
| French and Italian | 295 (32) | 155 (34) | 140 (30) | 1.17 (0.89-1.54) | 0.27 | 1.31 (0.98-1.75) | 0.07 |
| **Age at first neoplasm diagnosis** |  |  |  |  |  |  |  |
| Median (IQR, years) | 8.7 (3.0-13.4) | 8.7 (3.0-13.4) | 8.6 (3.6-13.3) |  |  |  |  |
| 0-4 years | 311 (34) | 166 (36) | 145 (31) | 1.14 (0.83 – 1.57) | 0.35 | 1.08 (0.75-1.56) | 0.4 |
| 5-9 years | 212 (23) | 97 (21) | 115 (25) | 0.84 (0.59 - 1.20) |  | 0.79 (0.54-1.14) |  |
| 10-14 years | 287 (31) | 144 (31) | 143 (31) | Ref. |  | Ref. |  |
| 15-20 years | 118 (13) | 56 (12) | 62 (13) | 0.90 (0.58 - 1.38) |  | 0.95 (0.61-1.48) |  |
| **Year of diagnosis** |  |  |  |  |  |  |  |
| 1976-1985 | 110 (12) | 54 (12) | 56 (12) | 0.84 (0.55 - 1.30) | **0.02** | ‡ |  |
| 1986-1995 | 193 (21) | 78 (17) | 115 (25) | 0.59 (0.41 - 0.85) |  | ‡ |  |
| 1996-2005 | 312 (34) | 164 (35) | 148 (32) | 0.97 (0.71 - 1.33) |  | ‡ |  |
| 2006-2016 | 313 (34) | 167 (36) | 146 (31) | Ref. |  | ‡ |  |

|  |  |  |  |  |  |
| --- | --- | --- | --- | --- | --- |
| **Diagnostic group of first primary neoplasm (ICCC-3)** |  |  |  |  |  |
| Leukaemias, myeloproliferative and myelodysplastic diseases | 35 (4) | 14 (3) | 21 (5) | 0.77 (0.37-1.58) | 0.38 |
| Lymphomas | 190 (21) | 95 (21) | 95 (20) | 1.15 (0.79-1.68) |  |
| CNS tumours and miscellaneous intracranial and intraspinal neoplasms | 256 (28) | 119 (26) | 137 (30) | Ref. |  |
| Neuroblastoma and other peripheral nervous cell tumours | 78 (8) | 45 (10) | 33 (7) | 1.57 (0.94-2.62) |  |
| Retinoblastoma | 44 (5) | 18 (4) | 26 (6) | 0.8 (0.42-1.53) |  |
| Renal tumours | 33 (4) | 20 (4) | 13 (3) | 1.77 (0.85-3.71) |  |
| Hepatic tumours | 24 (3) | 15 (3) | 9 (2) | 1.92 (0.81-4.54) |  |
| Malignant bone tumours | 100 (11) | 56 (12) | 44 (10) | 1.47 (0.92-2.33) |  |
| Soft tissue and other extraosseous sarcomas | 94 (10) | 48 (10) | 46 (10) | 1.2 (0.75-1.93) |  |
| Germ cell tumours, trophoblastic tumours, and neoplasms of gonads | 64 (7) | 29 (6) | 35 (8) | 0.95 (0.55-1.65) |  |
| Other malignant epithelial neoplasms and malignant melanomas | 6 (1) | 2 (<1) | 4 (1) | 0.58 (0.1-3.2) |  |
| Langerhans cell histiocytosis | 4 (<1) | 2 (<1) | 2 (<1) | 1.15 (0.16-8.3) |  |
| **Chemotherapy** |  |  |  |  |  |
| No | 61 (7) | 25 (5) | 36 (8) | Ref. |  |
| Yes | 867 (93) | 438 (95) | 429 (92) | 1.47 (0.87-2.49) | 0.15 |
| - With platinum agents | 491 (54) | 259 (56) | 232 (50) | 1.27 (0.98-1.64) | 0.07 |
| **Radiotherapy** |  |  |  |  |  |
| No | 319 (34) | 160 (35) | 159 (34) | Ref. |  |
| Yes | 609 (66) | 303 (65) | 306 (66) | 0.98 (0.75-1.29) | 0.91 |
| - With head radiotherapy (≥30 Gy) | 343 (37) | 166 (36) | 177 (38) | 0.91 (0.70-1.19) | 0.49 |
| - With chest radiotherapy | 328 (35) | 156 (34) | 172 (37) | 0.87 (0.66-1.33) | 0.29 |

**Haematopoietic stem cell transplantation**
|  |  |  |  |  |  |
| --- | --- | --- | --- | --- | --- |
| No | 864 (93) | 428 (92) | 436 (94) | Ref. |  |
| Yes | 64 (7) | 35 (8) | 29 (6) | 1.23 (0.74-2.05) | 0.43 |

**Relapse of first neoplasm**
|  |  |  |  |  |  |
| --- | --- | --- | --- | --- | --- |
| No | 738 (80) | 368 (80) | 370 (80) | Ref. |  |
| Yes | 190 (21) | 95 (21) | 95 (20) | 1.01 (0.73-1.38) | 0.97 |
| **Cancer predisposition syndrome** |  |  |  |  |  |  |  |
| No | 879 (95) | 442 (96) | 437 (94) | Ref. |  | Ref. |  |
| Yes | 49 (5) | 21 (5) | 28 (6) | 0.74 (0.42-1.33) | 0.31 | 0.54 (0.29-1.0) | 0.05 |
| **Second primary neoplasm** |  |  |  |  |  |  |  |
| No | 889 (96) | 438 (95) | 451 (97) | Ref. |  | Ref. |  |
| Yes | 39 (4) | 25 (5) | 14 (3) | 1.84 (0.94-3.58) | 0.07 | 1.92 (0.97-3.82) | 0.06 |
| **Age at study invitation** |  |  |  |  |  |  |  |
| Median (IQR, years) | 26.5 (18.8-36.5) | 25.1 (18.3-35.2) | 27.7 (19.5-37.3) |  |  |  |  |
| <10 years | 50 (5) | 23 (5) | 27 (6) | 0.72 (0.4-1.32) | **0.005** | 0.75 (0.39-1.43) | **0.002** |
| 10-19 years | 215 (23) | 121 (26) | 94 (20) | 1.09 (0.77-1.56) |  | 1.17 (0.81-1.70) |  |
| 20-29 years | 296 (32) | 160 (35) | 136 (29) | Ref. |  | Ref. |  |
| 30-39 years | 198 (21) | 78 (17) | 120 (26) | 0.55 (0.38-0.8) |  | 0.54 (0.38-0.79) |  |
| 40 or more years | 169 (18) | 81 (18) | 88 (19) | 0.78 (0.54-1.14) |  | 0.72 (0.48-1.07) |  |

**Legend:** ‡, omitted from multivariable regression model for collinearity with age at study invitation; CNS, central nervous system; ICC-3, international classification of childhood cancer, edition 3; IQR, interquartile range; n, number

**Figure 1.**
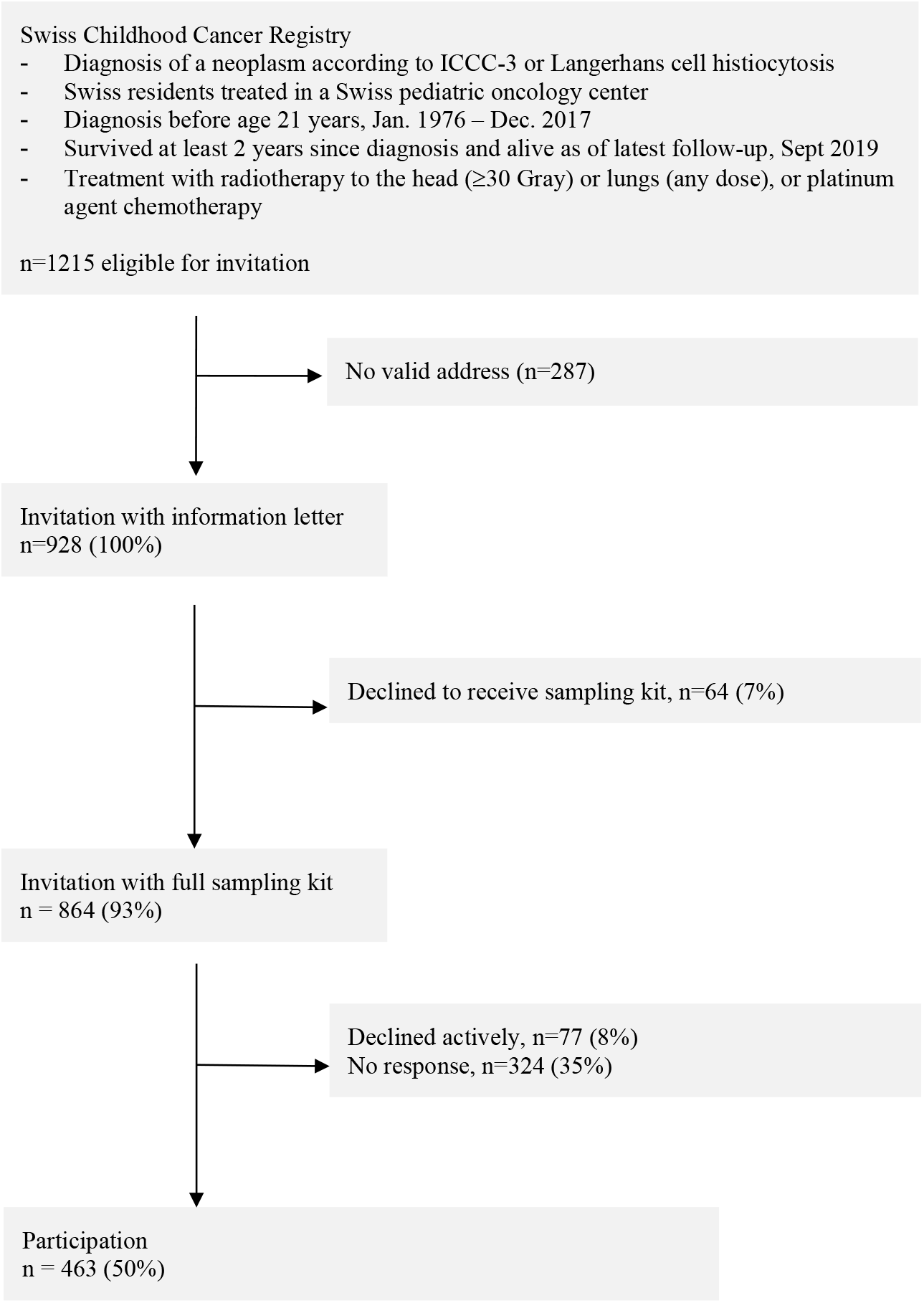
Flowchart of the response to invitation to the BISKIDS home collection of germline DNA in 928 patients. **Legend:** ICCC-3, international classification of childhood cancer, edition 3; n, number

### 3.2. Recruitment and impact of reminders

We invited 928 survivors for home saliva collection. After the first information letter, 64 (7%) actively declined participation through an opt-out reply form. The remaining 864 survivors received a saliva sampling kit and an informed consent form. Seventy-seven of those actively refused participation (8%) and 324 eligible survivors did not answer (35%). Within 6 weeks after the send out of the sampling kit, 291 (63%) participated (**Supplementary Figure 1**). The first reminder letter led to 117 additional participants (25%) and the second reminder to an additional 55 participants (12%). On the consent form, 383 (83%) indicated that they wanted to be informed about potential incidental findings and 394 (85%) agreed that their samples could be used after death.

### 3.3. Covariates associated with participation

Foreign nationality (odds ratio [OR] 0.5, 95%-confidence interval [CI] 0.4-0.7) and age at study were associated with non-participation. Survivors aged 30-39 years participated least (OR 0.5, CI 0.4-0.8), those 10-29 years tended to participate best, and those <10 years and >40 years in between. Those with a cancer predisposition syndrome tended to participate less (OR 0.5, CI 0.3-1.0), while survivors with a diagnosis of a second primary neoplasm (OR 1.9, CI 1.0-3.8) and those who spoke French or Italian (OR 1.3, CI 1.0-1.8) tended to participate more. There were no differences in sex, age at first primary neoplasm diagnosis, diagnostic groups, treatments, and relapse of first primary neoplasm between participants and non-participants.

## 4. DISCUSSION

This study described participation of Swiss childhood cancer survivors in home saliva collection for germline DNA extraction. We confirmed previous findings of about 50% participation in home DNA self-collection. Foreign nationality was a strong predictor for non-participation. Survivors with known CPSs and aged 30-39 years were also less likely to participate. In contrast, survivors with second primary neoplasms were more likely to participate. Most participating survivors wanted to be informed about incidental findings that might be relevant to their health and most agreed to their samples being available for research after their death.

The overall response rate of 50% was comparable to other DNA sample self-collections from home. Ness et al. reported 54% participation on 10,356 eligible adult childhood cancer survivors[5] and Dykema found 54% participation in 8,081 adults in the Wisconsin Longitudinal Study.[11] Nishita et al. offered an incentive of 25US$ for returning a saliva sample which led to 59% participation of 967 adults in a smoking cessation trial, which was only slightly higher than in studies without incentive.[12] Lower participation was found in genetic assessment of preterm birth where 23% of 708 mothers participated.[13] Higher participation rates were achieved in an active clinical setting with patient-caregiver interaction where 97% of 155 adults from the US participated.[14]

Foreign nationality was strongly associated with non-participation in concordance with a previous study from our group on response to questionnaire studies.[15] This could be related to language issues affecting understanding or reluctance to participate in research due to lower confidence in research. Foreign nationality is also linked to lower socioeconomic status, which was a predictor for lower participation in research.[16] Easy to understand information material, maybe in more languages, and information by phone might counteract these differences in participation. Our findings on language difference were in contrast to a large Swiss survey on willingness to participate in personalized health research,[16] and our group’s previous publication on participation in a questionnaire study,[15] where German speaking people were more likely to participate. The former was sent from the previously treating institutions and the latter form a German speaking research institution. In our study, the invitation was sent off from Geneva hospital, situated in the French speaking language region, which might have improved acceptance of the study of the CCS living in this language region. Survivors with known CPSs were less willing to participate in our study. Reasons could be the impression of futility as participants were already investigated, fear of further findings, or bad experiences with genetic workup. Survivors with second primary neoplasms tended to participate more, potentially because they were seeking explanations for their multiple neoplasms. Lower participation in older survivors might be reflective of being more distanced to the previous disease or in a busy phase of their lives (caring for children and being active at work).

A limitation of our study was the lack of information on reasons for non-participation. We further did not send additional sampling kits with reminders which might have prevented survivors from participating who had discarded their kit already. Strengths were the availability of clinical information including CPSs. Our results of the study are likely representative for childhood cancer survivors in other countries with similar trust in healthcare institutions and responsible (genetic) research.

Our study illustrates that home saliva collection through a process completely relying on mail is feasible and yields a response rate of about 50%. Specific survivor groups (foreign nationality, age 30-39 years, and patients with cancer predisposition syndromes) were less likely to participate which needs to be taken into account when studies are planned.

## Supporting information

Supplementary File. STROBE Checklist

## Data Availability

Datasets are available from the corresponding author on reasonable request for specific projects.

## Abbreviations table

BaHOP: Biobank for Hematology and Oncology in Pediatrics
BISKIDS: Germline DNA Biobank Switzerland for Childhood Cancer and Blood Disorders
CCS: Childhood cancer survivor
CNS: Central nervous system
CPS: Cancer predisposition syndrome
DNA: Deoxyribonucleic acid
GECCOS: Genetic risks for childhood cancer complications Switzerland
HSCT: Hematopoietic stem cell transplantation
ICCC-3: International Classification of Childhood Cancer, third edition
ISPM: Institute of Social and Preventive Medicine
SCCR: Swiss Childhood Cancer Registry
SPN: Second primary neoplasm

## Acknowledgements

We thank all childhood cancer patients, survivors, and families for participating in our study. We thank the study team of the Childhood Cancer Research Group, Institute of Social and Preventive Medicine, University of Bern, and SCCSS (Tomas Slama, Fabienne Luzi, Maria Otth, Selma Riedo), the data managers of the Swiss pediatric oncology clinics (Claudia Althaus, Nadine Assbichler, Pamela Balestra, Heike Baumeler, Nadine Beusch, Sarah Blanc, Pierluigi Brazzola, Susann Drerup, Janine Garibay, Franziska Hochreutener, Monika Imbach, Friedgard Julmy, Eléna Lemmel, Heike Markiewicz, Annette Reinberg, Renate Siegenthaler, Astrid Schiltknecht, Beate Schwenke, and Verena Stahel) and the data managers and administrative staff of the SCCR (Meltem Altun, Erika Brantschen, Katharina Flandera, Elisabeth Kiraly, Verena Pfeiffer, Julia Ruppel, Ursina Roder, and Nadine Lötscher). We thank the study team of the CANSEARCH research platform in paediatric oncology and haematology of the University of Geneva (Khalil Ben Hassine, Simona Jurkovic Mlakar, Vid Mlakar, Shannon Robin), and the Onco-Hematology Unit of the HUG (Frederic Baleydier, Fanette Bernard, André von Bueren, Fabienne Gumy-Pause, Nelly Hafner-Bénichou, Rodolfo Lo Piccolo).

## Competing interests

The authors declare that they have no competing interests

## STROBE Checklist

Supplementary File.

## Funding

This study is supported by the CANSEARCH Foundation for BISKIDS, the host biobank BaHOP, the research study GECCOS, and salary support to Nicolas Waespe and Sven Strebel. Further funding comes from the Swiss National Science Foundation (31BL30_185396), and Swiss Cancer Research (KFS-4722-02-2019, KLS/KFS-4825-01-2019).

The funding bodies have no role in the design of the study and collection, analysis, and interpretation of data and in writing this manuscript.

## Ethics approval and consent to participate

The Geneva Cantonal Commission for Research Ethics has approved the BaHOP biobank (approval PB_2017-00533) and the Genetic Risks for Childhood Cancer Complications Switzerland (GECCOS) study, which will utilize the samples (approval 2020-01723).

## Trial registration

Clinicaltrials.gov: NCT04702321

## Consent for publication

All participants signed informed consents themselves or through their guardians.

## Availability of data and materials

Datasets are available from the corresponding author on reasonable request for specific projects.

## Authors’ contributions

NW: Conceptualization, Design, Methodology, Formal analysis, Data curation, Project administration, Writing - all stages, Visualization.

SSt: Project administration, Writing – Reviewing and Editing

FB, LM: Writing – Reviewing and Editing

TN: Conceptualization, Design, Methodology, Writing – Reviewing and Editing

DM, VM: Conceptualization, Data curation, Project administration, Writing – Reviewing and Editing

FM: Data curation, Project administration, Visualisation, Writing – Reviewing and Editing

AS: Conceptualization, Design, Methodology, Writing - Reviewing and Editing.

CEK, MA: Supervision, Conceptualization, Design, Methodology, Writing - Reviewing and Editing.

All authors have approved the submitted version.

## Notes

### Competing Interest Statement

The authors have declared no competing interest.

### Clinical Trial

NCT04702321

