## Supplementary Figure 1 for "Predictors for participation in DNA self-sampling of childhood cancer survivors in Switzerland"

**Supplementary Figure 1.** Recruitment over time of Swiss childhood cancer survivors invited for home germline DNA collection.

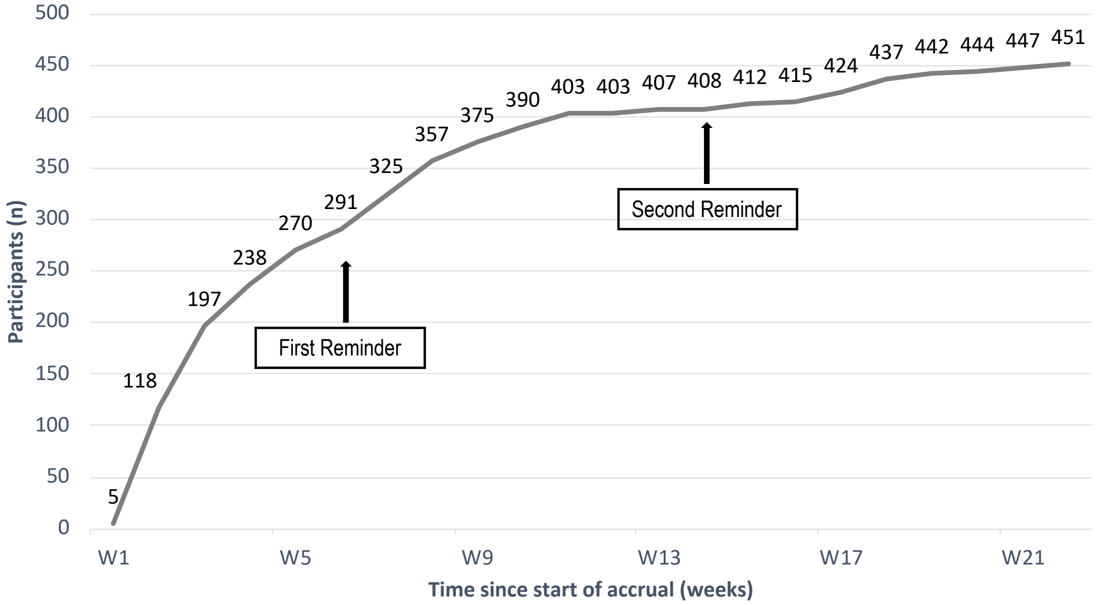


**Legend:** n, number; W, week.
